# Comparing health care use after hospital visit for SARS-CoV-2, respiratory syncytial virus, and other respiratory tract infections in children

**DOI:** 10.1101/2021.11.22.21266522

**Authors:** Vilde Bergstad Larsen, Ketil Størdal, Kjetil Telle, Fredrik Methi, Karin Magnusson

## Abstract

**Aim:** To explore whether children in specialist care with COVID-19 have increased post-discharge health care use when compared to children in specialist care with 1) respiratory syncytial virus (RSV) infection, and 2) other respiratory tract infections (RTIs).

**Methods:** In 34,214 children aged 1 month to 5 years who were registered with one or more hospital visit (outpatient or inpatient) with a diagnosis of COVID-19 (N=128), RSV infection (N=4,009), or other RTIs (N=34,458) from 2017-2021, we used a difference-in-differences study design to investigate the individual all-cause primary and specialist health care use from 12 weeks prior to 12 weeks after the hospital visit, stratified on infants (1-12 months) and children (1-5 years).

**Results:** We found a slight increase in primary health care use in the first four weeks after the hospital visit for infants with COVID-19 when compared to infants with RSV infection (6 [95%CI=2 to 13] per 10,000, 0.52% relative increase). For infants diagnosed with COVID-19, we found a similar post-visit increase in inpatients when compared to infants with RSV infection, which lasted for 12 weeks.

**Conclusions:** Our findings imply slightly increased health care use among infants after hospital visit for COVID-19 than among infants with other respiratory tract infections, for which potential etiological mechanisms deserve future clinical research. Severe COVID-19 in young children will not represent any markedly increased burden on the health services.

## Introduction

The infection prevention and control measures implemented to limit transmission of SARS-CoV-2 during 2020 and 2021 resulted in both a lower incidence of COVID-19 as well as lower circulation of other respiratory tract viruses.^1,2^ Currently, most disease control measures have been eased in Western countries due to high vaccine coverage. However, children under the age of five remain unvaccinated in most countries, and many may have a so-called immunity debt to other infections due to lack of viral exposure.^3,4^ Hence, the number of hospital contacts due to severe COVID-19, respiratory syncytial virus (RSV) infection, or other respiratory tract infections (RTIs) in Europe may be expected to increase.^5-7^

The extent to which children with severe COVID-19 have elevated health care use when compared to children severely ill with other viral infections, such as RSV infection or influenza, is unknown. Hospital care with RSV infection, which can be regarded as a proxy for severe RSV infection, has been reported to have a significant impact on post-disease health and morbidities in young children (caregivers report poorer health at 60 days after discharge)^8^, yet its impact when compared to the impact of COVID-19 has never been studied. Improved knowledge of how post-COVID health and health care use compares to post-RSV infection-, and post-RTI health and health care use, is important when developing and prioritizing vaccination of young children against RSV, influenza, and SARS-CoV-2.

Thus, we aimed to explore whether, and for how long, COVID-19 resulting in specialist care gives an increase in individual health care use in comparison to specialist care for RSV infection and other RTI in children aged 1-12 months and 1-5 years.

## Methods

Using a pre-post study design with two comparison groups, we utilized nation-wide individual-level registry data from the Norwegian Emergency Preparedness Register, Beredt C19 (Figure 1).^9^ The register covers all hospital visits (inpatient and outpatient), for all RTIs, including RSV, influenza, and SARS-CoV-2. Inpatient and outpatient care at Norwegian hospitals require referral from primary care, i.e., general practitioners or emergency wards, except for acute hospitalizations. All children admitted to Norwegian hospitals with RTI or symptoms of RTI are routinely tested for a panel of pathogens, including RSV and SARS-CoV-2 (the latter from March 2020). Medical diagnoses are coded based on pathogen testing and clinical presentation form in accordance with the International Classification of Diseases (ICD-10) system.

**Figure 1.**
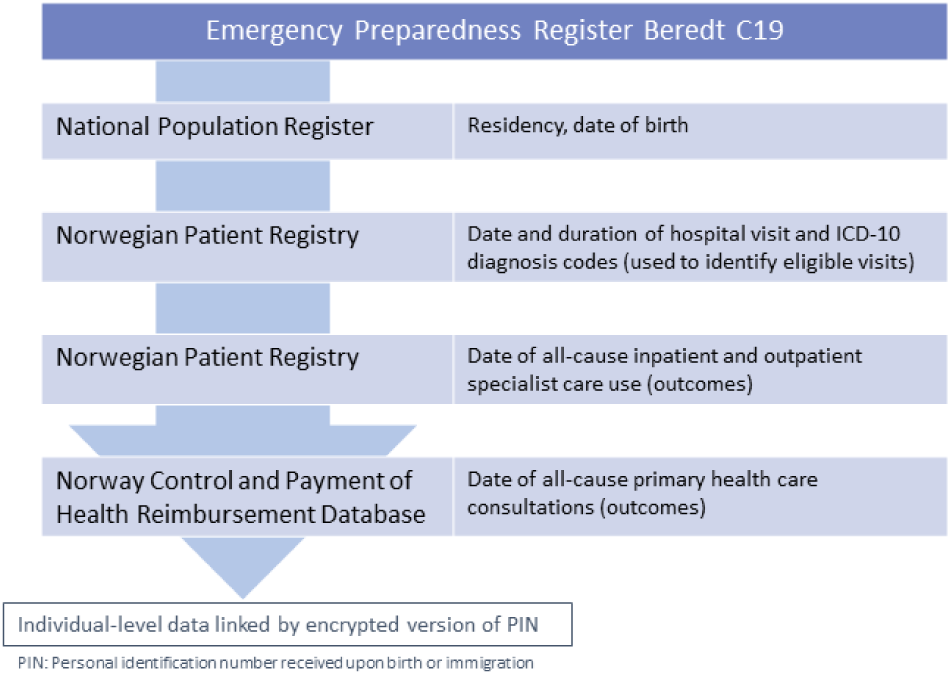
Description of registries from within Beredt C19 used to define study population and outcome variables.

### Study sample

Our study sample included all children aged 1 month to 5 years who were residents in Norway and who had a hospital visit with a diagnosis of COVID-19, RSV infection, or other RTI, between February 1^st^, 2017, and September 20^th^, 2021. The children with a hospital visit were assumed to have a more severe disease course than children without hospital visits, i.e., hospitalization was regarded as a proxy for severe disease. Eligible hospital visits were identified using ICD-10 diagnostic codes, categorized into three mutually exclusive diagnosis categories: COVID-19, RSV infection, and other RTI including influenza (both primary and secondary diagnoses included, see Table S1-1 for full list of included ICD-10 codes). To define our sample, we included both inpatients (overnight hospitalizations and in need for specialist care) and outpatients (not requiring overnight hospital stay yet in need for specialist care), independent of duration and urgency. We used the terms admission and discharge to describe the first and last day of a hospital visit. Hospital visits with any mention of the included diagnostic codes that occurred with less than two calendar days in between, were coded as one hospital visit, i.e., with the same hospital visit ID and earliest date of admission and latest date of discharge.

We divided our sample into two age groups (1-12 months and 1-5 years old at date of admission), because infected infants (1-12 months) are likely to have higher risk of hospital contact and post-disease health care use than older children (1-5 years).^10-12^ Children with more than one hospital visit during our study period reentered the study after a washout period of 6 months to allow for multiple hospital visits without conflating exposure with outcome.

Existing studies have reported relatively high rates of coinfection with SARS-CoV-2 and other respiratory pathogens among children.^13^ Coinfections (i.e., multiple mentions of different diagnosis categories within the same hospital visit) were very rare in our sample (1.4 % of all hospital visits). In these cases, we chose the most specific ICD-10 code (i.e., with known pathogen). No children in our sample had a hospital visit registered with both COVID-19 and RSV infection. However, our data did not contain information about testing for coinfections, and it is thus likely that many coinfections were missed.

### Outcomes

We studied all-cause primary health care use (including consultations at general practitioner and emergency ward) and all-cause specialist health care use (overall and separated into inpatient and outpatient services) before and after each child’s hospital visit for COVID-19, RSV infection, or other RTIs. We constructed our outcome data in a relative-week panel structure (relative to the admission date and the discharge date of the hospital visit). All outcomes were measured twelve weeks prior to hospital admission and twelve weeks after hospital discharge. All children contributed with a minimum of four weeks follow-up time before and after the hospital visit, allowing us to include newborn infants from age 1 month. More specifically, we followed each child from January 1st, 2017 (study start), date of birth, or date of immigration, whichever came last. Similarly, all children were followed until October 18th, 2021 (study end), until the day they turned 6 years old, death, or emigration, whichever came first. With a maximum observation period of twelve weeks before and twelve weeks after the hospital visit for both age groups, we obtained panel data that were equally balanced (i.e., the similar amount of relative week data for both age groups allowed for valid comparison across the age groups).

### Statistical analysis

First, we presented descriptive statistics on our study sample, as well as the crude rates of primary and specialist health care use before and after the hospital visit (calculated as the weekly percentage of children with at least one primary/specialist consultation), by age and diagnosis group. To estimate how much more, or less, frequent utilization of health care services was after the hospital contact for children with COVID-19 compared to 1) children with RSV infection, and 2) children with other RTIs, we used a difference-in-differences (DiD) design with separate estimates for each four-week period after hospital discharge. This allowed us to isolate the impact of being diagnosed with COVID-19 from being diagnosed with RSV infection or other RTIs on post-disease health care use. Hence, we extended the traditional DiD design from one to three post-treatment periods: 1-4 weeks; 5-8 weeks; and 9-12 weeks after discharge, compared to the baseline twelve weeks prior to admission. The statistical models were estimated using linear regression with robust standard errors on the individual level and adjusting for calendar month and year to account for any seasonal variation in health care utilization.^14-17^

To assess whether results were driven by changed patterns of health care use and health-seeking behavior due to the pandemic, we conducted a post-hoc sensitivity analysis for each outcome including only hospital contacts from March 12^th^, 2020, when national COVID-19 lockdown measures were first announced in Norway, to September 20^th^, 2021, using the same approach as described above (see Supplementary material 2 for further details).

In total, only 25 children aged 18 and younger were registered with codes indicative of Multiorgan Inflammatory Syndrome after COVID-19 (MIS-C) in Norway by June 2021, and no further analyses of this subgroup was undertaken due to the even lower number that can be expected when restricting to our age group of interest.^18^

Statistical software R v. 4.0.2 was used for data preparation and statistical analyses.

### Ethics

The establishment of an emergency preparedness register forms part of the legally mandated responsibilities of The Norwegian Institute of Public Health (NIPH) during epidemics. Institutional board review was conducted, and The Ethics Committee of South-East Norway confirmed (June 4^th^, 2020, #153204) that external ethical board review was not required.

## Results

Of a total of 653,544 children born after February 1^st^, 2011, we identified 34,214 children fulfilling our inclusion criteria (Figure 2). A total of 4,840 hospital visits were excluded due to insufficient follow-up time (Figure 2). Among 12,058 children aged 1-11 months and 23,682 children aged 1-5 years who were registered with 38,594 hospital visits in the period from February 1^st^, 2017, to September 20^th^, 2021, we studied primary and specialist health care use before and after 128 hospital visits for COVID-19 (among 53 children aged 1-11 months and 75 children aged 1-5 years), 4,009 hospital visits for RSV infection (among 2,438 children aged 1-11 months and 1,571 children aged 1-5 years), and 34,457 hospital visits for other RTIs (among 9,695 children aged 1-11 months and 24,762 children aged 1-5 years) (Table S1-2). Children with RSV infection had somewhat longer average length of stay than children with COVID-19 and other RTI (Table S1-2).

**Figure 2.**
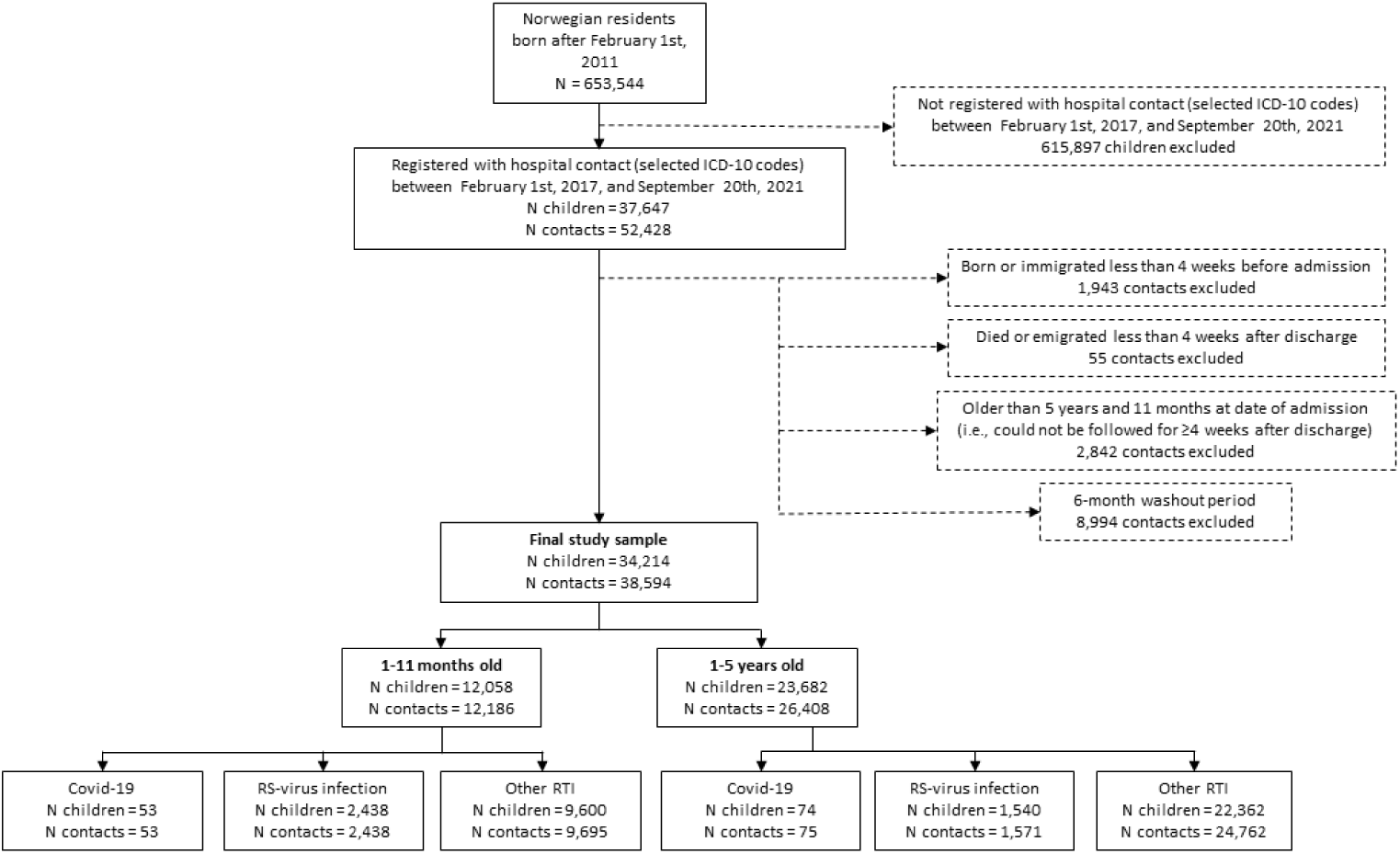
Flowchart of study population.

### Crude all-cause primary and specialist health care use

Infants aged 1-11 months had similar levels of primary health care use in the twelve weeks prior to the hospital visit for all diagnoses, while pre-admission use of primary care was slightly higher for children aged 1-5 years with RSV infection than for same-aged children with COVID-19 or other RTIs (Table S1-3). The lowest average level of primary health care use prior to the hospital visit was found for children aged 1-5 years with COVID-19 (Table S1-3). Use of primary health care increased for all diagnosis groups and both age groups in the seven days before the hospital visit (Figure 3). This is to be expected, because hospital care depends on referral from primary care at least when the degree of urgency is low. Within the first four weeks of discharge, children with RSV infection and other RTIs had resumed to pre-admission levels of primary care use, while it remained elevated until weeks 5-8 for the children with COVID-19 (Figure 3; Table S1-3).

**Figure 3.**
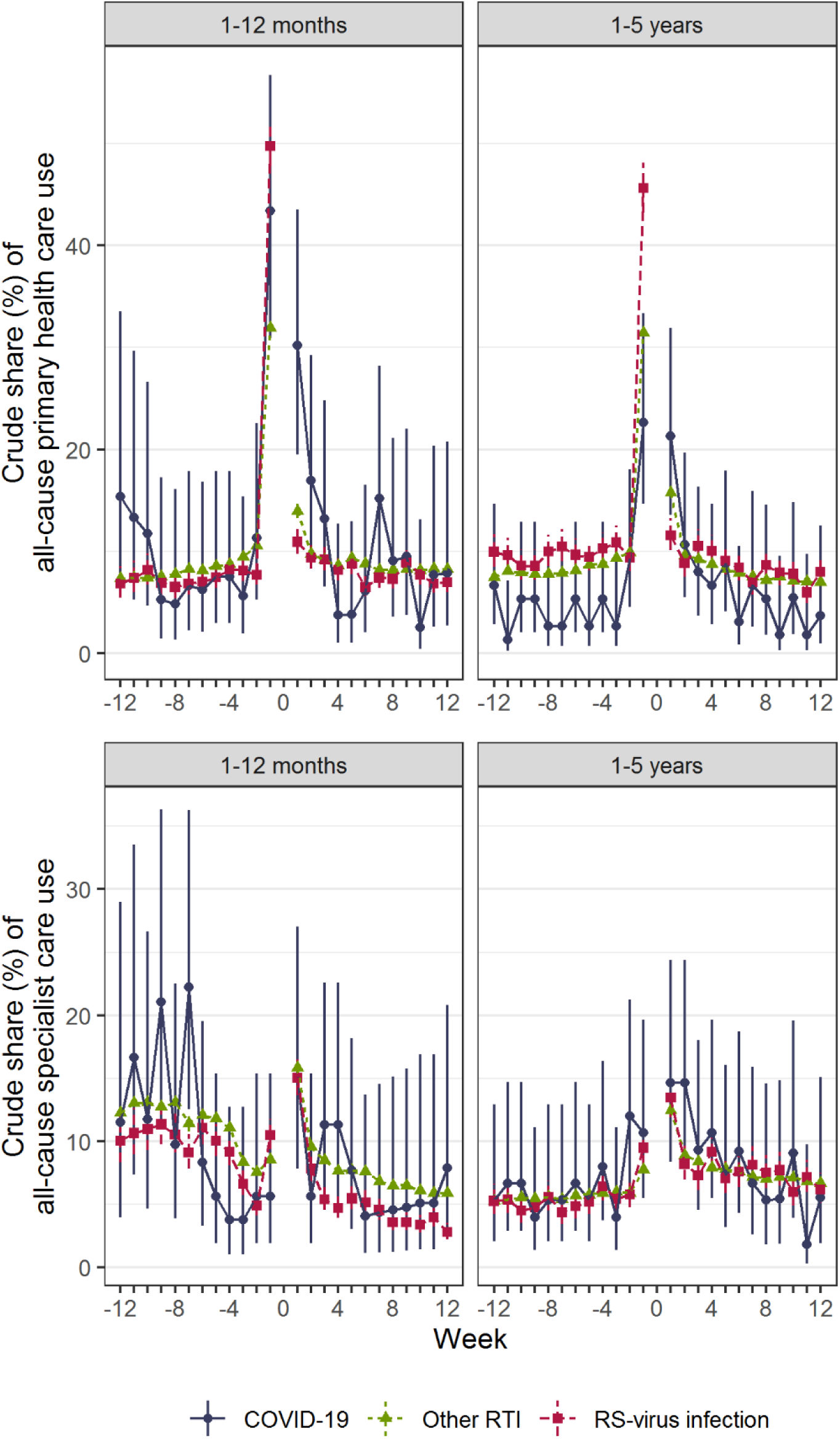
Crude share (%) of primary and specialist health care use in the weeks before and after hospitalization. Figure 3 note: 95 % confidence intervals (CI) calculated using the Wilson method for binomial proportions. For each week (7-day period) before and after hospitalization, the denominator equaled the number of children who could be observed the given week and the numerator equaled the number of children who had at least one health care contact (calculated separately for age and diagnosis groups).

The average weekly utilization of specialist care prior to admission was similar to the level of primary health care use for all groups, except for children aged 1-5 years with RSV infection or other RTI (Figure 3; Table S1-3). For most children, we observed an increase in the crude percentage that utilized specialist health care the week after discharge (Figure 3). Like primary health care use, utilization of specialist health care resumed to pre-admission levels within 5-8 weeks of discharge for most diagnosis and age groups (Table S1-3).

### Differences-in-differences analyses, all-cause primary care use

Children aged 1-12 months with COVID-19 had a short-term increase in primary care use after the hospital visit compared to same-aged children with RSV infection (0.064 percentage points, 0.52 % relative increase), corresponding to an average weekly increase of approximately 6 consultations per 10,000 children with a hospital visit for COVID-19 (Table 1). Similarly, children aged 1-5 years also had an increase in utilization of primary health care the first four weeks after discharge, when compared to children with RSV infection (0.068 percentage points, 0.53 % relative increase) or other RTIs (0.046 percentage points, 0.45 % relative increase) (Table 1). These differences corresponded to average weekly increases of approximately 7 and 5 consultations per 10,000 children with COVID-19, respectively. After 5 weeks of discharge, there were no differences in utilization of primary health care for either age group. The elevated primary care use among infants was confirmed in our sensitivity analysis (0.086 percentage points, 0.73% relative increase), where we restricted the comparison groups to children with hospital contact during the COVID-19 pandemic (Table S2-1). We found no differences in primary care use for children aged 1-5 years in the sensitivity analysis.

**Table 1.** Impacts of hospital visit for COVID-19 on primary and specialist health care use in children, using children with hospital visit for RSV infection and other RTIs as comparison group.

|  | Week 1-4 |  | Week 5-8 |  | Week 9-12 |  |
| --- | --- | --- | --- | --- | --- | --- |
| Primary health care use | $\beta$ | 95 % CI | $\beta$ | 95 % CI | $\beta$ | 95 % CI |
| **1-11 months** |  |  |  |  |  |  |
| RSV infection | 0.064* | 0.002, 0.126 | -0.001 | -0.065, 0.062 | -0.019 | -0.078, 0.04 |
| Other RTI | 0.044 | -0.013, 0.101 | -0.019 | -0.077, 0.04 | -0.025 | -0.084, 0.033 |
| **1-5 years** |  |  |  |  |  |  |
| RSV infection | 0.068* | 0.022, 0.144 | 0.034 | -0.007, 0.076 | 0.016 | -0.019, 0.05 |
| Other RTI | 0.046* | 0.002, 0.091 | 0.022 | -0.019, 0.063 | 0.003 | -0.027, 0.032 |
| **Specialist health care use** | $\beta$ | 95 % CI | $\beta$ | 95 % CI | $\beta$ | 95 % CI |
| **1-12 months** |  |  |  |  |  |  |
| RSV infection | 0.041 | -0.002, 0.084 | 0.014 | -0.047, 0.074 | 0.028 | -0.041, 0.097 |
| Other RTI | 0.02 | -0.021, 0.06 | -0.006 | -0.057, 0.045 | 0.011 | -0.046, 0.067 |
| **1-5 years** |  |  |  |  |  |  |
| RSV infection | 0.005 | -0.037, 0.047 | -0.018 | -0.07, 0.034 | -0.014 | -0.062, 0.034 |
| Other RTI | 0.014 | -0.026, 0.054 | -0.017 | -0.072, 0.038 | -0.025 | -0.069, 0.02 |

Notes: Differences-in-differences estimates ( $\beta$ , in percentage points) for the change in the percentage of children
\*Statistically significant on 5 % level.

### Differences-in-differences analyses, all-cause specialist care use

Children with COVID-19 had no increased use of all-cause specialist care services (inpatients and outpatients studied together) after discharge when compared to children with RSV infections or other RTIs (Table 1). However, when restricting to use of inpatient services, we observed that children with COVID-19 had an increase in post-discharge utilization both for weeks 1-4 (0.044 percentage points, 0.95 % relative increase), 5-8 (0.037 percentage points, 0.80 % relative increase) and 9-12 (0.038 percentage points, 0.82 % relative increase) when compared to children with RSV infection, yet only for the youngest children (1-12 months) (Table S1-4). This corresponds to a weekly average increase of approximately 4 inpatients per 10,000 children. No increase was observed for outpatient care (Table S1-4). These findings were largely confirmed in our sensitivity analyses (Tables S2-1, S2-2). In addition, the sensitivity analyses showed elevated use of specialist care for infants aged 1-12 months with COVID-19 when compared to children with RSV infection (0.096 percentage points, 0.83% relative increase) (Table S2-1). When studying inpatient care during the pandemic, the differences in utilization after COVID-19 and RSV infection hospital visit among infants dissipated after 8 weeks, but we also found slightly elevated use of inpatient care when comparing hospital contact for COVID-19 to other RTIs in weeks 1-4 (0.029 percentage points, 0.45% relative increase) and in weeks 9-12 (0.035 percentage points, 0.54% relative increase) (Table S2-2).

## Discussion

In the current study of 34,214 children aged 1 month to 5 years, we found a minor increase in primary health care use in the first four weeks after hospital visit with COVID-19 diagnosis for infants aged 1-11 months, compared to infants with RSV infection, and for children aged 1-5 years, compared to same-aged children with RSV infection and other RTIs. This difference dissipated after five weeks of discharge. Further, infants with a hospital visit for COVID-19 had an increase of similar magnitude in use of inpatient care when compared to 1-11-month-olds with RSV infection, lasting for 12 weeks after discharge. The observed patterns of post-discharge health care use remained largely similar when restricting the study period to the pandemic (i.e., starting March 12^th^, 2020) in separate sensitivity analyses.

To our knowledge, the present study is the first to compare the major RTIs in terms of post-disease health care use. By including hospital visits for both the novel SARS-CoV-2 virus, RS virus, and other RT viruses including influenza, our findings form a significant contribution to the understanding of the burden of common RTIs on the health care services. An additional strength of our study is the utilization of routinely collected individual-level data from nation-wide registers that are mandated by law, ensuring representativeness and no attrition. Relatedly, our use of two comparison groups (RSV infection and other RTIs) as well as several post-discharge time periods strengthen the robustness of our findings.

### Comparison to previous studies

Few similar previous studies exist that allow for a valid comparison of findings. However, in a study of caregivers/parents reports about children’s health (age 30 months or less) following RSV discharge (N=45), poorer health was observed for up to 60 days after discharge when compared to age-matched controls (N=46).^8^ Here, we could compare the rather serious hospital visit for RSV with hospital visit for COVID-19 and other RTIs in 34,214 children aged 0-5 years using health care records. In this way, we could put the consequences of RSV infection and COVID-19 into context, showing they may be more or less equally severe in terms of post-discharge health and health care use, despite that the average duration of the initial visit was longer for children with RSV infection, indicating more severe disease. The slight increase in primary health care use after hospital visit for COVID-19 compared to after RSV infection or other RTIs is in line with results from our recent study showing that children with mild COVID-19 had increased utilization of primary health care after infection, when compared to same-aged non-infected children.^14^ The findings in these two studies do not necessarily indicate that children with COVID-19 have more severe post-disease symptoms than children with or without other diagnoses. Rather, it may reflect differences in health-seeking behavior or the behavior of health care services when facing the new SARS-CoV-2 virus compared to the more well-known infections due to e.g., RSV. Still, health care use may be regarded as a proxy for poor health. Although we did not aim to shed light on long-term complaints or clinical consequences in any of these studies, the etiological mechanisms for potentially worse complaints and/or more health-seeking behavior following COVID-19 among children than among healthy children or children ill with other diseases, deserve further research attention in clinical studies.

### Clinical implications

Our study is representative for all countries with equal access to health care services. Because hospital visits for RSV infection and other RTIs is considerably more common than hospital visits for COVID-19 among young children, our observations indicate that targeted interventions against RSV and other RTI should receive priority over COVID-19 when it comes to reducing the burden on the health services. Furthermore, there is evidence that the more contagious Omicron variant of the SARS-CoV-2 virus leads to milder disease than previous virus variants, also among children.^19-21^ Whether our findings, which are based on the Wuhan, Alpha, and Delta virus variants, over- or underestimates, or correctly estimates, post-discharge health care use following COVID-19 in children should thus be addressed in future studies.

### Limitations

Some limitations to our data and results should be noted. First, COVID-19 requiring inpatient or outpatient specialist care is extremely rare in children, which is reflected in the sample size and statistical strength even in our nation-wide population. Along this line, due to the suppressed incidence of hospital contacts for RSV and other RTI because of disease control measures during 2020 and 2021, we compared the incidence of the very few hospital contacts for COVID-19 during these years, with the much higher incidence of RSV infection and other RTI during 2017-2019. We handled this potential limitation by controlling for seasonal variation (calendar month and year) in our models and by running sensitivity analyses where we included RSV and other RTI hospital contacts only from March 2020 and onwards, with similar findings. Our similar observations across the main and sensitivity analyses may indicate that potential violations to the common trend assumption was not an issue in our study. Thus, we regard any potential difference in health-seeking behavior prior to and after the implementation of lockdown restrictions in March 2020 as unlikely to have impacted on our findings, at least among infants. A second limitation may be that for children in most age and diagnosis groups, we found similar utilization levels for specialist health care as for primary health care in the weeks prior to hospital admission. This may indicate that children requiring hospital care due to the included diseases generally have poorer health and thus different patterns of health-seeking behavior than the general population. Thus, hospital care may not serve as a proxy for disease severity, but rather reflect the underlying health of the child. However, both inpatient and outpatient hospital contact generally require referral from a general practitioner or emergency ward. We therefore studied those potentially more severely ill than infected children not in contact with any health care services or only in contact with primary care, as they were judged by a physician to require specialist examination or treatment. Lastly, our 12-week observation period may be too short to capture post-viral conditions, such as RSV followed by repeated episodes of bronchopulmonal obstructions.

Finally, there was no general guideline on mandatory post-COVID follow-up examinations in Norway, as has been performed in some studies.^22,23^ However, we cannot exclude that the small increase in care visits observed for children hospitalized with COVID-19 was due to the caregivers requesting a closer follow-up, which was not requested for children hospitalized with other RTI.

## Conclusion

Our findings imply a slight increase in post-discharge health care use among children with a hospital diagnosis of COVID-19 than among children with hospital diagnoses of other RTIs. However, hospital visits among children with COVID-19 was rare and the increase in post-COVID health-care use was small, implying that the potential increase in health care use post-COVID compared to post-RSV or post-RTI will not represent any markedly increased or lasting burden on the health services. If health care contacts are regarded as proxies for poor health, we believe that the etiological mechanisms for potentially worse complaints following COVID-19 than following other RTIs in children should be further explored.

## Supporting information

Supplementary material 2 - sensitivity analyses

## Data Availability

The individual-level data used in this study are not publicly available due to privacy laws, but the individual-level data in the registries compiled in Beredt C19 are accessible to authorized researchers after ethical approval and application to helsedata.no administered by the Norwegian Directorate of eHealth.

## Data sharing statement

The individual-level data used in this study are not publicly available due to privacy laws, but the data in the registries compiled in Beredt C19 are accessible to authorized researchers after ethical approval and application to helsedata.no administered by the Norwegian Directorate of eHealth.

