## Supplementary material 1 for "Comparing health care use after hospital visit for SARS-CoV-2, respiratory syncytial virus, and other respiratory tract infections in children"

**Table S1-1:** Selected International Classification of Diseases (ICD-10) codes and diagnosis categories.

| Category | ICD-10 codes |
|---|---|
| COVID-19 | U07.1 U07.2 |
| RSV infection | J12.1 J20.5 J21.0 |
| Other RTI | J00 J02.0 J02.8 J02.9 J03.0 J03.8 J03.9 J04.0 J04.1 J04.2 J05.0 J05.1 J06.0 J06.8 J06.9 J12.0 J12.2 J12.3 J12.8 J12.9 J13 J14 J15.0 J15.1 J15.2 J15.3 J15.4 J15.5 J15.6 J15.7 J15.8 J15.9 J16.0 J16.8 J17.0 J17.1 J17.2 J17.3 J17.8 J18.0 J18.1 J18.2 J18.8 J18.9 J22 J20.0 J20.1 J20.2 J20.3 J20.4 J20.6 J20.7 J20.8 J20.9 J09 J10.0 J10.1 J10.8 J11.0 J11.1 J11.8 |

**Table S1-2:** Descriptive statistics.

| | 1-12 months | | | 1-5 years | | |
|---|---|---|---|---|---|---|
| | **COVID-19** | **RSV** | **Other RTI** | **COVID-19** | **RSV** | **Other RTI** |
| Children, N | 34,214 | | | | | |
| Children, N | 12,058 | | | 23,682 | | |
| Children, N | 53 | 2,438 | 9,600 | 74 | 1,540 | 22,362 |
| Contacts, N | 53 | 2,438 | 9,695 | 75 | 1,571 | 24,762 |
| Person-weeks, N | 1,141 | 53,346 | 220,617 | 1,742 | 37,824 | 607,009 |
| Females, N (%) | 23 (43.4) | 1,072 (44.0) | 3,975 (41.0) | 33 (44.0) | 670 (42.6) | 10,313 (41.6) |
| Age, median [25%-75%] | 2 [2-4] | 2 [1-5] | 3 [2-7] | 2.4 [1.55-3.5] | 1.6 [1.2-2.3] | 2.1 [1.4-3.2] |
| Avg. length of stay, days (SD) | 1.43 (3.98) | 3.40 (3.62) | 1.05 (5.02) | 1.15 (1.67) | 3.11 (4.31) | 0.93 (2.53) |

Note: Age presented as median and interquartile range (IQR) age in months/years at admission. The calculation of age and sex percentages are done using the number of unique hospital visits in the denominator. Due to the 6-month washout period, some children appear in more than one of the age and/or diagnosis groups.

**Table S1-3:** Crude percentages of primary and specialist health care use by age and diagnosis group.

| | 1-12 months | | | | | | 1-5 years | | | | | |
|---|---|---|---|---|---|---|---|---|---|---|---|---|
| | COVID-19 | | RSV | | Other RTI | | COVID-19 | | RSV | | Other RTI | |
| Week | P | S | P | S | P | S | P | S | P | S | P | S |
| Pre | 11.76 | 9.68 | 12.21 | 9.26 | 10.83 | 10.98 | 6.00 | 6.67 | 12.95 | 5.64 | 10.33 | 5.82 |
| 1-4 | 16.04 | 10.85 | 9.44 | 8.25 | 10.39 | 10.3 | 11.67 | 12.33 | 10.26 | 9.55 | 10.85 | 9.45 |
| 5-8 | 8.38 | 5.24 | 7.52 | 4.69 | 8.65 | 7.16 | 6.02 | 7.23 | 8.29 | 7.57 | 7.74 | 7.41 |
| 9-12 | 6.96 | 5.70 | 7.62 | 3.45 | 8.26 | 6.10 | 3.21 | 5.50 | 7.42 | 6.76 | 7.19 | 6.97 |

Note: Weekly percentages of children utilizing primary (P) and specialist (S) health care in the 12-week pre-admission period (pre) and 1-4-, 5-8-, and 9-12-week post-discharge period.

**Table S1-4:** Impacts of hospital visit for COVID-19 on outpatient and inpatient specialist health care use in children, using children hospitalized with RSV infection and other RTI as comparison group.

| | Week 1-4 | | Week 5-8 | | Week 9-12 | |
|---|---|---|---|---|---|---|
| **Outpatient care use** | **β** | **95 % CI** | **β** | **95 % CI** | **β** | **95 % CI** |
| **1-11 months** | | | | | | |
| RSV infection | 0.025 | -0.023-0.072 | -0.008 | -0.076-0.06 | 0.012 | -0.062-0.086 |
| Other RTI | 0.017 | -0.025-0.06 | -0.015 | -0.073-0.044 | 0.002 | -0.062-0.065 |
| **1-5 years** | | | | | | |
| RSV infection | 0 | -0.035-0.035 | -0.019 | -0.061-0.023 | -0.018 | -0.062-0.026 |
| Other RTI | 0.006 | -0.028-0.04 | -0.021 | -0.064-0.022 | -0.028 | -0.066-0.011 |
| **Inpatient care use** | **β** | **95 % CI** | **β** | **95 % CI** | **β** | **95 % CI** |
| **1-12 months** | | | | | | |
| RSV infection | 0.044* | 0.012-0.076 | 0.037* | 0.004-0.069 | 0.038* | 0.001-0.076 |
| Other RTI | 0.021 | -0.005-0.048 | 0.019 | -0.007-0.044 | 0.03 | -0.002-0.062 |
| **1-5 years** | | | | | | |
| RSV infection | 0.002 | -0.029-0.033 | -0.001 | -0.033-0.031 | -0.006 | -0.035-0.023 |
| Other RTI | 0.002 | -0.028-0.033 | 0.001 | -0.033-0.035 | -0.007 | -0.033-0.02 |

Notes: Differences-in-differences estimates (β, in percentage points) for the change in the percentage of children utilizing outpatient and inpatient specialist health care per week after the day of discharge. The 12-week pre-admission period for the comparison group (RSV infection or other RTI) is used as reference category. Average outpatient care in the 12-week pre-admission period by age and diagnosis group is given in Table S1-5.
*Statistically significant on 5 % level.

**Table S1-5:** Crude percentages of outpatient and inpatient specialist care use by age and diagnosis group.

| | 1-12 months | | | | | | 1-5 years | | | | | |
|---|---|---|---|---|---|---|---|---|---|---|---|---|
| | COVID-19 | | RSV | | Other RTI | | COVID-19 | | RSV | | Other RTI | |
| Week | O | I | O | I | O | I | O | I | O | I | O | I |
| Pre | 6.83 | 4.36 | 5.51 | 4.62 | 6.69 | 5.50 | 5.89 | 1.67 | 4.67 | 1.17 | 4.85 | 1.34 |
| 1-4 | 9.43 | 4.25 | 6.51 | 2.03 | 7.85 | 3.36 | 9.67 | 3.33 | 7.77 | 2.31 | 7.48 | 2.62 |
| 5-8 | 4.19 | 2.62 | 3.66 | 1.16 | 5.62 | 2.09 | 5.62 | 2.41 | 5.86 | 2.03 | 6.10 | 1.83 |
| 9-12 | 5.06 | 3.16 | 2.66 | 0.88 | 4.87 | 1.62 | 4.13 | 1.38 | 5.44 | 1.56 | 5.82 | 1.59 |

Note: Weekly percentages of children utilizing outpatient (O) and inpatient (I) specialist health care in the 12-week pre-admission period (pre) and 1-4-, 5-8-, and 9-12-week post-discharge period.
