## Supplementary material 2 - sensitivity analyses for "Comparing health care use after hospital visit for SARS-CoV-2, respiratory syncytial virus, and other respiratory tract infections in children"

### Description of sensitivity analyses

To assess whether our results were driven by different patterns in health-seeking behavior due to the COVID-19 pandemic, we performed separate sensitivity analyses where we excluded all hospital visits (outpatient and inpatient) before March 12$^{th}$, 2020, when the first national lockdown measures were announced in Norway.

Our sensitivity analyses included 6,253 hospital visits among 1,700 children aged 1-12 months and 4,381 children aged 1-5 years in the period from March 12th, 2020, to September 20th, 2021. We studied utilization of primary and specialist health care use after 128 hospital visits for COVID-19; 144 hospital visits for RSV infection; and 5,981 hospital visits for other RTIs.

### Results

As in our main analysis, infants aged 1-12 months with COVID-19 had elevated use of primary health care services (0.086 percentage points, 0.73 % relative increase) lasting for 1-4 weeks after the hospital visit, compared to primary care use after hospital visit for RSV infection among infants (Table S2-1). An increase in specialist health care use (0.096 percentage points, 0.83 % relative increase) in the first four weeks after hospital contact was also observed when comparing infants aged 1-11 months with COVID-19 to same-aged infants with RSV infection. No differences in primary or specialist care after hospital visit for COVID-19 compared to RSV infection or other RTIs among children aged 1-5 years were found.

Infants aged 1-11 months also had increased utilization of inpatient services for 1-4 weeks (0.072 percentage points, 1.65% relative increase) and 5-8 weeks (0.07 percentage points, 1.61 % relative increase) after hospital visit for COVID-19 compared to after hospital visit for RSV infection (Table S2-2). The same pattern was observed when comparing infants aged 1-11 months with COVID-19 to infants with other RTIs, but only in weeks 1-4 (0.029 percentage points, 0.45% relative increase) and weeks 9-12 (0.035 percentage points, 0.54% relative increase). Similar to our main analysis, we found no differences in outpatient care use.

**Table S2-1:** Impacts of hospital visit for COVID-19 on primary and specialist health care use in children, using children with hospital visit for RSV infection and other RTIs as comparison group, restricted to hospital visits between March 12th, 2020, and September 20th, 2021.

| | Week 1-4 | | Week 5-8 | | Week 9-12 | |
|---|---|---|---|---|---|---|
| **Primary health care use** | **β** | **95 % CI** | **β** | **95 % CI** | **β** | **95 % CI** |
| **1-11 months** | | | | | | |
| RSV infection | 0.086* | 0.005, 0.167 | 0.036 | -0.046, 0.118 | 0.034 | -0.056, 0.125 |
| Other RTI | 0.05 | -0.007, 0.107 | -0.021 | -0.08, 0.038 | -0.021 | -0.08, 0.038 |
| **1-5 years** | | | | | | |
| RSV infection | 0.049 | -0.028, 0.127 | 0.008 | -0.062, 0.078 | -0.023 | -0.104, 0.058 |
| Other RTI | 0.043 | -0.001, 0.088 | 0.016 | -0.026, 0.057 | -0.003 | -0.033, 0.028 |
| **Specialist health care use** | **β** | **95 % CI** | **β** | **95 % CI** | **β** | **95 % CI** |
| **1-12 months** | | | | | | |
| RSV infection | 0.096* | 0.022, 0.169 | 0.072 | -0.029, 0.174 | 0.09 | -0.024, 0.204 |
| Other RTI | 0.026 | -0.016, 0.068 | -0.004 | -0.055, 0.048 | 0.014 | -0.043, 0.071 |
| **1-5 years** | | | | | | |
| RSV infection | 0.024 | -0.04, 0.087 | -0.034 | -0.116, 0.047 | -0.062 | -0.16, 0.036 |
| Other RTI | 0.01 | -0.021, 0.059 | -0.017 | -0.072, 0.039 | -0.024 | -0.069, 0.02 |

Notes: Differences-in-differences estimates (β, in percentage points) for the change in the percentage of children utilizing primary or specialist health care per week after the day of discharge. The 12-week pre-admission period for the comparison group (RSV infection or other RTI) is used as reference category. Average primary care use in the 12-week pre-admission period by age and diagnosis group is given in Table S2-3. *Statistically significant on 5 % level.

**Table S2-2:** Impacts of hospital visit for COVID-19 on outpatient and inpatient specialist health care use in children, using children with hospital visit for RSV infection and other RTIs as comparison group, restricted to hospital visits between March 12th, 2020, and September 20th, 2021.

| | Week 1-4 | | Week 5-8 | | Week 9-12 | |
|---|---|---|---|---|---|---|
| **Outpatient care use** | **β** | **95 % CI** | **β** | **95 % CI** | **β** | **95 % CI** |
| **1-11 months** | | | | | | |
| RSV infection | 0.025 | -0.023, 0.072 | -0.008 | -0.076, 0.06 | 0.012 | -0.062, 0.086 |
| Other RTI | 0.023 | -0.02, 0.066 | -0.01 | -0.069, 0.049 | 0.01 | -0.054, 0.073 |
| **1-5 years** | | | | | | |
| RSV infection | 0.036 | -0.015, 0.087 | -0.044 | -0.124, 0.036 | -0.047 | -0.146, 0.052 |
| Other RTI | 0.009 | -0.025, 0.043 | -0.022 | -0.065, 0.021 | -0.029 | -0.068, 0.009 |
| **Inpatient care use** | **β** | **95 % CI** | **β** | **95 % CI** | **β** | **95 % CI** |
| **1-12 months** | | | | | | |
| RSV infection | 0.072* | 0.02, 0.124 | 0.07* | 0.014, 0.126 | 0.066 | -0.001, 0.132 |
| Other RTI | 0.029* | 0.001, 0.057 | 0.024 | -0.002, 0.051 | 0.035* | 0.002, 0.068 |
| **1-5 years** | | | | | | |
| RSV infection | -0.026 | -0.078, 0.031 | -0.013 | -0.055, 0.028 | -0.025 | -0.065, 0.015 |
| Other RTI | 0 | -0.031, 0.031 | 0.001 | -0.033, 0.035 | -0.006 | -0.033, 0.021 |

Notes: Differences-in-differences estimates (β, in percentage points) for the change in the percentage of children utilizing outpatient and inpatient specialist health care per week after the day of discharge. The 12-week pre-admission period for the comparison group (RSV infection or other RTI) is used as reference category. Average outpatient care in the 12-week pre-admission period by age and diagnosis group is given in Table S2-4. *Statistically significant on 5 % level.

**Table S2-3:** Crude percentages of primary and specialist health care use by age and diagnosis group.

| | 1-12 months | | | | | | 1-5 years | | | | | |
|---|---|---|---|---|---|---|---|---|---|---|---|---|
| | COVID-19 | | RSV | | Other RTI | | COVID-19 | | RSV | | Other RTI | |
| Week | P | S | P | S | P | S | P | S | P | S | P | S |
| Pre | 12.8 | 9.68 | 11.8 | 11.6 | 9.87 | 12.9 | 6.0 | 6.67 | 10.3 | 8.38 | 8.06 | 6.72 |
| 1-4 | 16.0 | 10.8 | 8.33 | 6.41 | 8.91 | 11.5 | 11.7 | 12.3 | 10.6 | 11.7 | 9.23 | 10.1 |
| 5-8 | 8.38 | 5.24 | 4.69 | 3.12 | 8.07 | 8.81 | 6.02 | 7.23 | 7.69 | 13.8 | 6.6 | 8.76 |
| 9-12 | 6.96 | 5.7 | 6.25 | 1.34 | 7.28 | 7.7 | 3.21 | 5.5 | 7.43 | 14.2 | 6.01 | 8.31 |

Note: Weekly percentages of children utilizing primary (P) and specialist (S) health care in the 12-week pre-admission period (pre) and 1-4-, 5-8-, and 9-12-week post-discharge period.

**Table S2-4:** Crude percentages of outpatient and inpatient specialist health care use by age and diagnosis group.

| | 1-12 months | | | | | | 1-5 years | | | | | |
|---|---|---|---|---|---|---|---|---|---|---|---|---|
| | COVID-19 | | RSV | | Other RTI | | COVID-19 | | RSV | | Other RTI | |
| Week | O | I | O | I | O | I | O | I | O | I | O | I |
| Pre | 6.83 | 6.66 | 7.93 | 4.36 | 9.19 | 6.5 | 5.89 | 1.67 | 8.25 | ** | 6.03 | 1.39 |
| 1-4 | 9.43 | 4.25 | 5.45 | 1.6 | 9.73 | 3.36 | 9.67 | 3.33 | 7.95 | 4.92 | 8.52 | 2.99 |
| 5-8 | 4.19 | 2.62 | 3.12 | ** | 7.73 | 2.25 | 5.62 | 2.41 | 13.8 | 2.56 | 7.76 | 2.04 |
| 9-12 | 5.06 | 3.16 | ** | ** | 6.67 | 1.89 | 4.13 | ** | 13.5 | ** | 7.52 | 1.74 |

Note: Weekly percentages of children utilizing inpatient (I) and outpatient (O) health care in the 12-week pre-admission period (pre) and 1-4-, 5-8-, and 9-12-week post-discharge period. ** Not presented due to small numbers (<5) in numerator.
